# Zero-Shot Evaluation of Kimi K2 on Pediatric Clinical Cases

**DOI:** 10.1101/2025.07.29.25332368

**Authors:** Gianluca Mondillo, Mariapia Masino, Simone Colosimo, Alessandra Perrotta, Vittoria Frattolillo, Fabio Giovanni Abbate

## Abstract

**Background:** The application of large language models (LLMs) in pediatric medicine requires rigorous performance evaluation prior to clinical implementation.

**Objective:** To evaluate the accuracy of the Kimi K2 model in analyzing pediatric clinical cases using a zero-shot approach. Methods: 2,249 multiple-choice questions from pediatric clinical cases, ranging in age from 1 day to 16 years, extracted from the MedQA dataset were analyzed. The model was tested via API with standardized parameters, temperature set to zero, and zero-shot prompts. Accuracy was calculated by comparing the responses with the dataset’s ground truth.

**Results:** Kimi K2 achieved an overall accuracy of 78.39%, corresponding to 1,763 correct answers out of 2,249 total, with 100% of responses in the required format. Conclusions: The model demonstrates competitive performance for medical education and diagnostic support, while still having limitations that require human clinical supervision.

## 1 Introduction

The integration of Large Language Models (LLMs) into the medical field has opened new frontiers for diagnostic support and clinical education, demonstrating promising capabilities in analyzing complex cases and generating differential diagnoses [1]. However, despite significant progress documented in general medicine, the specific application of these tools in pediatrics remains a largely unexplored territory, characterized by a scarcity of dedicated studies and systematic clinical validations [2]. Pediatric medicine indeed presents unique challenges for artificial intelligence systems, primarily due to the intrinsic complexity of clinical presentations in developmental age, which require a sophisticated understanding of developmental and growth processes, as well as highly specialized knowledge of pediatric age-specific pathological patterns [2]. This specificity necessitates a rigorous evaluation of the performance of advanced language models before any consideration for clinical implementation. The MedQA dataset currently represents the most consolidated reference standard for the systematic evaluation of language models’ clinical competencies, incorporating a large corpus of multiple-choice questions derived from the USMLE (United States Medical Licensing Examination), which test in-depth medical knowledge and complex clinical reasoning abilities [3, 4]. In this context, Kimi K2, developed by Moonshot AI, emerges as an innovative technological solution in language model architecture. It is a highly sophisticated mixture-of-experts system that implements 384 specialized experts, using an architecture that selectively activates 32 billion parameters out of a total of 1 trillion, representing a significant advancement in computational optimization and inference performance [5]. Particularly relevant for clinical evaluation is the adoption of a zero-shot approach, a methodology that dispenses with the use of specific training examples for the target task, thus more faithfully simulating real operational conditions in which clinicians face new cases without analogous precedents [6]. This approach allows for a more ecologically valid evaluation of the model’s capabilities in authentic clinical scenarios. The primary objective of this study is therefore to conduct a systematic and rigorous evaluation of Kimi K2’s diagnostic accuracy in the analysis of pediatric clinical cases, using a zero-shot methodology to determine the model’s application potential as a support tool in contemporary pediatric practice.

## 2 Materials and Methods

2,249 multiple-choice questions extracted from the MedQA dataset were analyzed; inclusion criteria exclusively comprised pediatric clinical cases with patients aged between 1 day and 16 years and verified ground truth, each with at least four answer options. The sample composition is summarized in Table 1, which reports the number of questions for each of the five age bands considered.

**Table 1.**
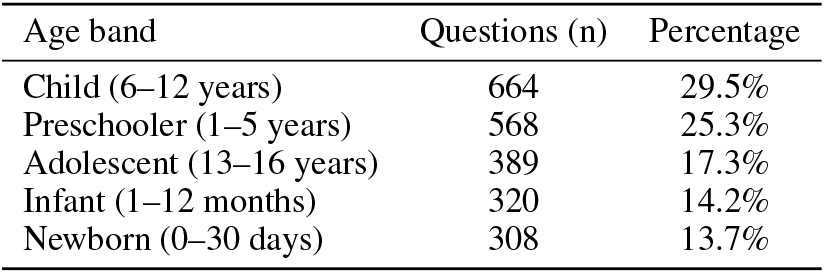
Distribution of the 2,249 questions by patient age band.

The Kimi K2 model was queried via Application Programming Interface (API), setting the temperature to zero to maximize reproducibility and adopting a zero-shot approach with sequential question presentation. The standardized prompt read: «Analyze the following pediatric clinical case and select the correct answer. Respond ONLY with the letter (A, B, C, D, or E) of the answer you deem correct.» followed by the case text and answer options. The generated responses were automatically compared with the ground truth, and accuracy was calculated as the percentage ratio of correct to total responses. Finally, a stratification by specialty was performed: for each clinical area, the number of questions, correct answers, specific accuracy, and balanced accuracy were calculated, the latter obtained as an unweighted average of the accuracies of the individual specialties, to provide a global measure independent of the numerical imbalance between categories.

## 3 Results

Out of 2,249 questions analyzed, Kimi K2 achieved an accuracy of 78.39%, corresponding to 1,763 correct answers (Figure 1). The model demonstrated complete reliability in the output format with 100% of responses automatically parsable, eliminating the need for manual post-processing. The total time required for the analysis was approximately 2 hours.

**Figure 1.**
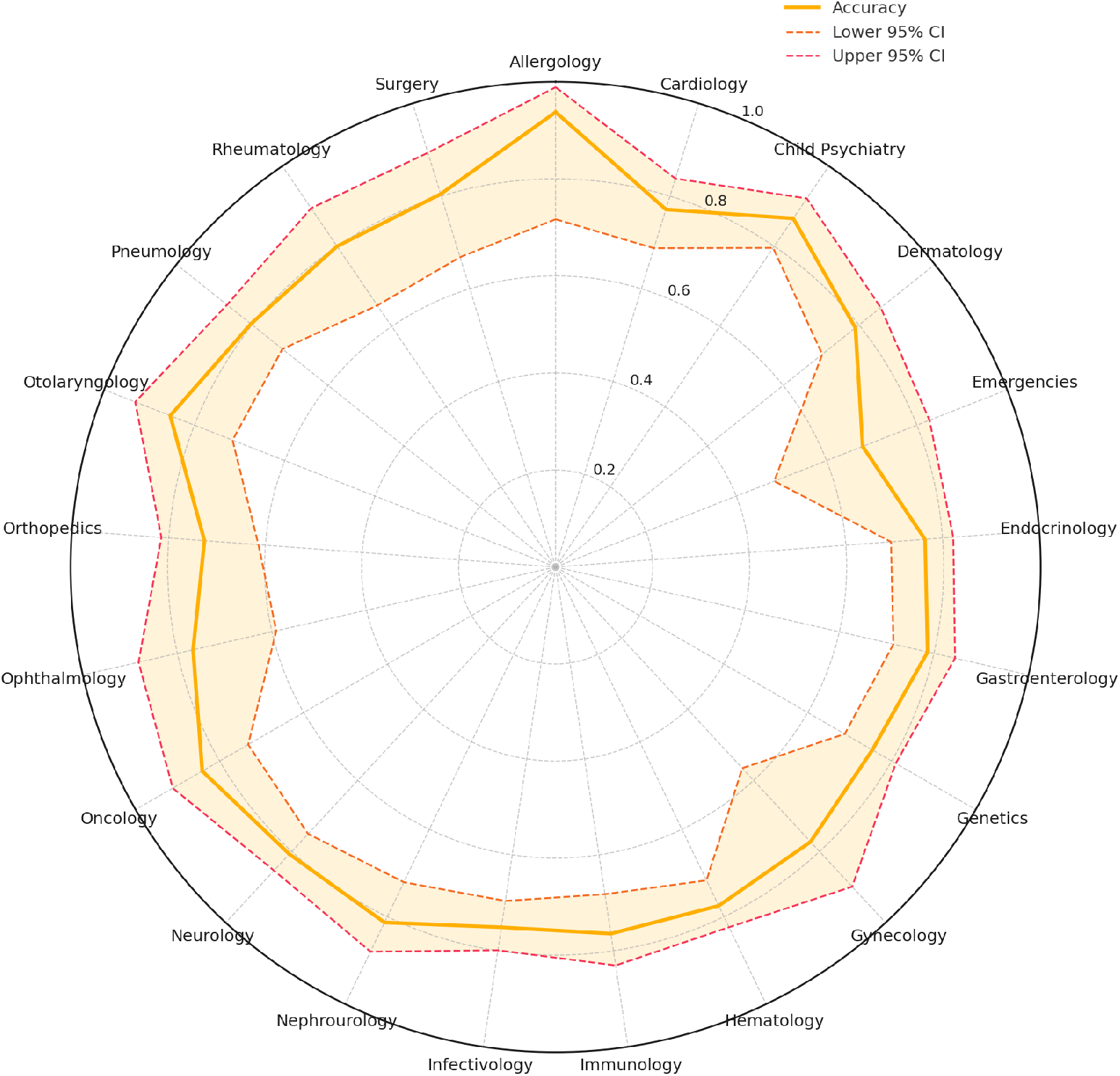
Radar chart of model accuracy by paediatric specialty. The solid yellow line depicts Kimi K2’s raw accuracy for each specialty, while the shaded orange band shows the 95 % Wilson confidence interval, bounded by the dashed red lines (lower limit) and dotted red lines (upper limit).

Specialty stratification revealed substantial variability in performance across paediatric clinical areas. As reported in Table 2, the distribution of questions across specialties was highly imbalanced. Results are summarised with a radar chart that, for each specialty *i*, shows the raw accuracy 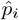 together with its 95% confidence interval estimated via the Wilson score method (Eq. 1). As expected, the interval width increases as the sample size *n*_*i*_ decreases, indicating greater statistical uncertainty for less represented specialties.

**Table 2.** Specialty Raw Accuracy with 95% Wilson Confidence Interval.

| Specialty | Questions (n) | Correct | Accuracy (%) | 95% CI |
| --- | --- | --- | --- | --- |
| Infectivology | 269 | 202 | 75.1 | 69.6–79.9 |
| Hematology | 231 | 179 | 77.5 | 71.7–82.4 |
| Neurology | 223 | 180 | 80.7 | 75.0–85.4 |
| Genetics | 191 | 144 | 75.4 | 68.8–81.0 |
| Endocrinology | 165 | 126 | 76.4 | 69.3–82.2 |
| Gastroenterology | 150 | 118 | 78.7 | 71.4–84.5 |
| Immunology | 123 | 94 | 76.4 | 68.2–83.1 |
| Cardiology | 118 | 91 | 77.1 | 68.8–83.8 |
| Child Psychiatry | 115 | 100 | 87.0 | 79.6–91.9 |
| Pneumology | 112 | 90 | 80.4 | 72.0–86.7 |
| Dermatology | 105 | 83 | 79.0 | 70.3–85.7 |
| Nephrourology | 91 | 74 | 81.3 | 72.1–88.0 |
| Orthopedics | 73 | 53 | 72.6 | 61.4–81.5 |
| Oncology | 63 | 53 | 84.1 | 73.2–91.1 |
| Surgery | 46 | 37 | 80.4 | 66.8–89.3 |
| Otolaryngology | 41 | 35 | 85.4 | 71.6–93.1 |
| Rheumatology | 40 | 32 | 80.0 | 65.2–89.5 |
| Ophthalmology | 30 | 23 | 76.7 | 59.1–88.2 |
| Emergencies | 25 | 17 | 68.0 | 48.4–82.8 |
| Gynecology | 22 | 17 | 77.3 | 56.6–89.9 |
| Allergology | 16 | 15 | 93.8 | 71.7–98.9 |

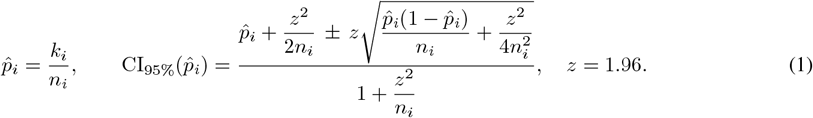

Here *k*_*i*_ is the number of correct answers and *n*_*i*_ the total number of questions in specialty *i*; *z* = *z*_1−*α*/2_ is the standard normal quantile for the desired confidence level (1.96 for 95%).

Additionally, we computed the balanced accuracy (Eq. 2), a global metric not influenced by the numerical imbalance between categories; in our data it equals 79.20%.

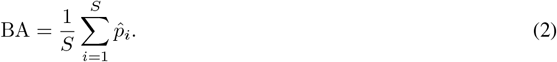

where *S* is the total number of specialties considered.

Table 2 presents the distribution of questions by specialty and, for each, the accuracy with its 95% confidence interval calculated according to Wilson’s method.

Figure 2 visualizes the percentage distribution of questions by age band and specialty, highlighting the dataset’s imbalance along these two dimensions.

**Figure 2.**
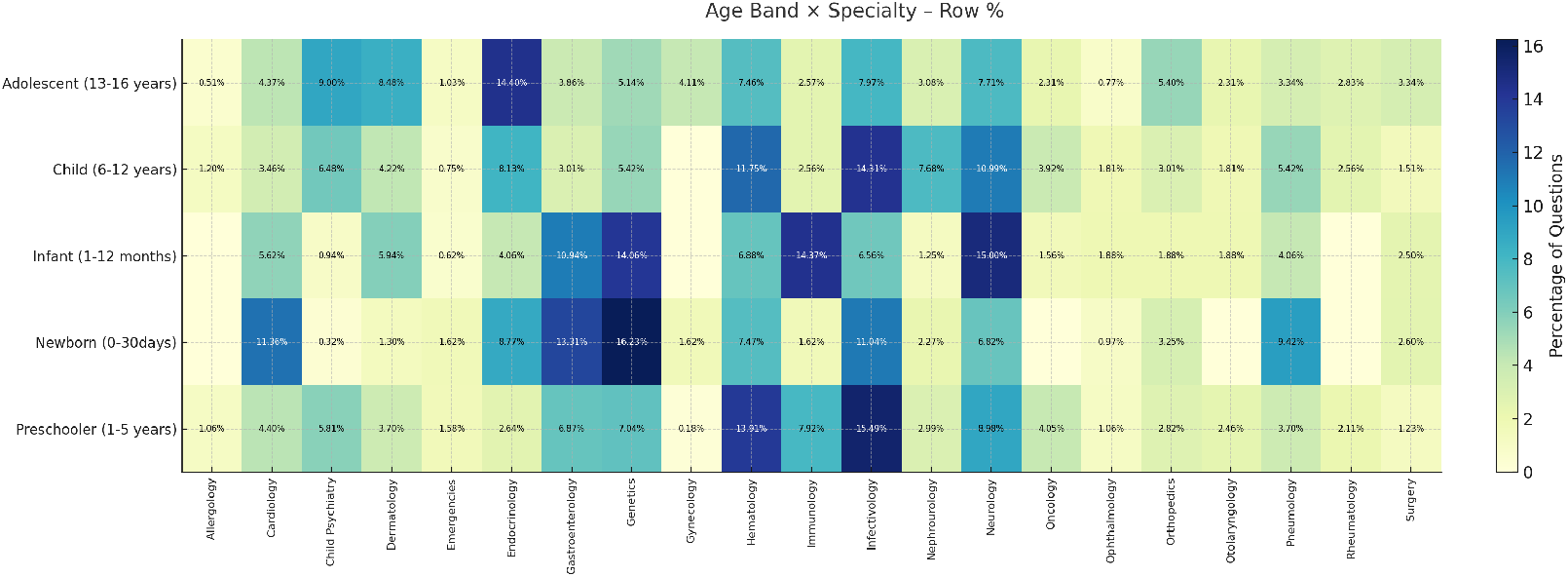
Heat map showing the percentage distribution of the 2,249 multiple choice questions across patient age bands and paediatric specialties. Within each row the values sum to 100%; thus, each cell represents the share of questions in that age band that falls under the corresponding specialty. Darker shades highlight combinations that are more prevalent in the dataset, whereas lighter cells indicate under represented areas.

## 4 Discussion

Kimi K2’s accuracy of 78.39% compares favorably to previous studies on LLMs in pediatric medicine. GPT-3.5 achieved 60.2% on MedQA-USMLE [7], ChatGPT 74.5% on pediatric emergency medicine [8], while ChatGPT-4 only 39% on complex cases from the New England Journal of Medicine [9]. However, advanced reasoning models such as ChatGPT O1 with 92.8% and DeepSeek-R1 with 87.0% significantly outperform Kimi K2 [10]. This difference is attributable to Kimi K2’s “non-thinking” nature, which does not use explicit chain-of-thought reasoning but integrates implicit planning as a reflexive agentic model. Kimi K2 exhibits notable technical performance within the landscape of advanced language models, with a Measuring Massive Multitask Language Understanding (MMLU) score of 0.824 and an Intelligence Index of 58 [11]. MMLU is a benchmark that evaluates general knowledge across 57 academic disciplines, while the Intelligence Index represents an aggregated measure of the model’s cognitive abilities. On specific benchmarks, the model achieves 87.8% on standard MMLU and 69.2% on MMLU-Pro, a more challenging version of the test, demonstrating excellence even in specialized domains such as GPQA-Diamond, which includes advanced university-level questions in physics, chemistry, and biology [12]. However, it is important to emphasize that these general benchmark performances do not necessarily predict accuracy in specific medical domains, as evidenced by the gap between the model’s general capabilities and the results obtained on pediatric clinical cases in this study. Figure 2 also highlights an age imbalance in the dataset: over half of the questions concern school-aged and preschool children, while newborns, infants, and adolescents are much less represented. This distribution may have favored the model in disciplines typical of early life stages and limited its performance in specialties whose complexity primarily emerges during puberty. Future studies should therefore balance the sample by age band to more accurately evaluate the model’s reliability across the entire spectrum of pediatric development. Kimi K2’s mixture-of-experts architecture represents a significant advancement in computational efficiency [5]. The model utilizes 384 specialized experts with selective activation of 32 billion parameters out of 1 trillion total [5], optimizing computational resources while maintaining high performance. This architecture allows for achieving state-of-the-art performance in advanced knowledge, mathematics, and coding among non-thinking models [5]. The implementation of the MuonClip optimizer constitutes a critical technical breakthrough: it enabled stable training of a trillion-parameter model “with zero training instability” [13], overcoming the traditional limitations of optimizers like AdamW on such large scales. The zero-shot approach used reflects an increasingly recognized methodology for evaluating LLMs in medical contexts, particularly relevant when specific training data are limited [6]. This approach allows for a more ecologically valid evaluation of the model’s capabilities in authentic clinical scenarios, more faithfully simulating the real operational conditions in which clinicians face new cases without analogous precedents [6]. An accuracy exceeding 65%, which represents the minimum threshold for medical certifications [8], suggests potential applications in medical education and specialized training [14, 15], in support for differential diagnoses [16, 17], in pediatric telemedicine assistance [18, 19], and in the review of complex clinical cases [9, 16]. The model’s agentic capabilities, defined as the ability to autonomously use external tools, execute code, and complete complex multi-step tasks without human intervention [5], open possibilities for advanced decision support systems that could assist clinicians in analyzing complex cases through automated workflows integrating data from multiple clinical sources. The main limitations include an error rate of 22% which limits autonomous use in critical contexts [1, 9], the lack of explainability with an absence of explicit clinical reasoning [9, 6], generalizability issues as benchmark performance is not necessarily translatable to clinical practice [1, 10], and limited validation requiring prospective studies on real cases [1]. The model presents limitations in termini of speed of 44.3 tokens per second, which is below average, and a context window size of 130k tokens, which is smaller than other advanced models [11], potentially affecting the analysis of extensive clinical documentation. From the perspective of clinical implementation, the two available versions, Kimi-K2-Base for research and customization and Kimi-K2-Instruct for chat and agentic applications [5], offer flexibility for various healthcare needs. The availability of APIs compatible with OpenAI/Anthropic standards facilitates integration into existing healthcare systems [5], while the possibility of fine-tuning the base model could allow for specific specializations in pediatric subdisciplines. The need for human medical supervision remains fundamental to ensure safety in clinical implementation [20].

## 5 Conclusions

Kimi K2 demonstrates competitive performance in the analysis of pediatric clinical cases with a zero-shot approach, achieving an accuracy of 78.58%. These results support the model’s potential as an educational and clinical support tool, while still necessitating human medical supervision. Future research should focus on pediatric-specific fine-tuning, on the implementation of explainability systems, on prospective validation in real clinical contexts, and on the development of safety frameworks for clinical implementation. The model represents a significant step forward towards the integration of AI in pediatrics, yet requiring further development before widespread clinical adoption.

## Data Availability

All data produced in the present study are available upon reasonable request to the authors

## References

[1] Singhal, K and others. Large language models encode clinical knowledge. Nature, 620(7972):172–180, 2023. doi: 10.1038/s41586-023-06291-2. Epub 2023 Jul 12. Erratum in: Nature. 2023 Aug;620(7973):E19. doi: 10.1038/s41586-023-06455-0.

[2] Mondillo, G and others. Artificial Intelligence in Pediatrics: An Opportunity to Lead, not to Follow. J Pediatr, 283:114641, 2025. doi: 10.1016/j.jpeds.2025.114641.

[3] Jin, D., Pan, E., Oufattole, N., Weng, W.-H., Fang, H., & Szolovits, P. What Disease Does This Patient Have? A Large-Scale Open Domain Question Answering Dataset from Medical Exams. Applied Sciences, 11(14):6421, 2021. 10.3390/app11146421.

[4] Kaggle. MedQA-USMLE: A Large-scale Open Domain Question Answering Dataset from Medical Exams. 2025. https://www.kaggle.com/datasets/moaaztameer/medqa-usmle. Accessed 19 July 2025.

[5] Moonshot AI Team. Kimi K2: Open Agentic Intelligence. GitHub Repository. 2025. https://github.com/MoonshotAI/Kimi-K2. Accessed 19 July 2025.

[6] Kojima, Takeshi & Gu, Shixiang & Reid, Machel & Matsuo, Yutaka & Iwasawa, Yusuke. Large Language Models are Zero-Shot Reasoners. 2022. 10.48550/arXiv.2205.11916.

[7] Liévin, V, Hother, CE, Motzfeldt, AG, Winther, O. Can large language models reason about medical questions? Patterns (N Y), 5(3):100943, 2024. doi: 10.1016/j.patter.2024.100943.

[8] Ramgopal S, Varma S, Gorski JK, Kester KM, Shieh A, Suresh S. Evaluation of a Large Language Model on the American Academy of Pediatrics’ PREP Emergency Medicine Question Bank. Pediatric Emergency Care. 2024 Dec 1;40(12):871–875. doi: 10.1097/PEC.0000000000003271.

[9] Barile, J and others. Diagnostic Accuracy of a Large Language Model in Pediatric Case Studies. JAMA Pediatrics, 178(3):313–315, 2024. doi: 10.1001/jamapediatrics.2023.5750.

[10] Mondillo, G and others. Comparative Evaluation of Advanced AI Reasoning Models in Pediatric Clinical Decision Support: ChatGPT O1 vs. DeepSeek-R1. medRxiv. 2025. doi: 10.1101/2025.01.27.25321169

[11] Artificial Analysis. Kimi K2 - Intelligence, Performance & Price Analysis. 2025. Disponibile: https://artificialanalysis.ai/models/kimi-k2.

[12] Gupta, M. Kimi-k2 Benchmarks explained. Medium. 2025. https://medium.com/data-science-in-your-pocket/kimi-k2-benchmarks-explained-5b25dd6d3a3e. Accessed 25 July 2025.

[13] Moonshot AI Team. Kimi K2: Open Agentic Intelligence Technical Report Of KIMI K2 Technical Documentation. 2025. https://github.com/MoonshotAI/Kimi-K2/blob/main/tech_report.pdf. Accessed 25 July 2025.

[14] Suresh, S and Misra, SM. Large Language Models in Pediatric Education: Current Uses and Future Potential. Pediatrics, 154(3):e2023064683, 2024. doi: 10.1542/peds.2023-064683.

[15] Abd-Alrazaq, A, AlSaad, R, Alhuwail, D, Ahmed, A, Healy, PM, Latifi, S, Aziz, S, Damseh, R, Alabed Alrazak, S, Sheikh, J. Large Language Models in Medical Education: Opportunities, Challenges, and Future Directions. JMIR Med Educ, 9:e48291, 2023. doi: 10.2196/48291.

[16] McDuff, D, Schaekermann, M, Tu, T, et al. Towards accurate differential diagnosis with large language models. Nature, 642:451–457, 2025. 10.1038/s41586-025-08869-4.

[17] Young, CC, Enichen, E, Rivera, C, Auger, CA, Grant, N, Rao, A, Succi, MD. Diagnostic Accuracy of a Custom Large Language Model on Rare Pediatric Disease Case Reports. Am J Med Genet A, 197(2):e63878, 2025. doi: 10.1002/ajmg.a.63878.

[18] John Snow Labs. Large Language Models in Telemedicine and Remote Care. 2025. Disponibile: https://www.johnsnowlabs.com/large-language-models-in-telemedicine-and-remote-care/. Accessed 26 July 2025.

[19] Burke, BL Jr, Hall, RW; SECTION ON TELEHEALTH CARE. Telemedicine: Pediatric Applications. Pediatrics, 136(1):e293–308, 2015. doi: 10.1542/peds.2015-1517.

[20] Harrison, S, Despotou, G, Arvanitis, TN. Hazards for the Implementation and Use of Artificial Intelligence Enabled Digital Health Interventions, a UK Perspective. Stud Health Technol Inform, 289:14–17, 2022. doi: 10.3233/SHTI210847.

